# Review of publicly available state reimbursement policies for removal and reinsertion of Long-Acting Reversible Contraception

**DOI:** 10.1101/2024.05.10.24307204

**Authors:** Ekwutosi M. Okoroh, Charlan D. Kroelinger, Olivia R. Sappenfield, Julia F. Howland, Lisa M. Romero, Keriann Uesugi, Shanna Cox

## Abstract

We examined reimbursement policies for the removal and reinsertion of long-acting reversible contraception (LARC). We conducted a standardized, web-based review of publicly available state policies for language on reimbursement of LARC removal and reinsertion. We also summarized policy language on barriers to reimbursement for LARC removal and reinsertion. Twenty-six (52%) of the 50 states had publicly available policies that addressed reimbursement for LARC removal. Of these, 14 (28%) included language on reimbursement for LARC reinsertion. Eleven states included language on barriers for reimbursement for removal and/or reinsertion: five state policies included language with other requirements for removal only, three policies included language with additional requirements for reinsertion only, and three included language with additional requirements for both. Three state policies specified no barriers be placed on reimbursement for removal and one specified no barriers be placed on reimbursement for reinsertion. Half of the states in the U.S. do not have publicly available policies on reimbursement for the removal and reinsertion of LARC devices. Inclusion of unrestricted access to these services is important for reproductive autonomy.

## Introduction

Reproductive autonomy includes the right to decide and control contraceptive use [1–2]. Long-acting reversible contraception (LARC) methods (i.e. defined as intrauterine devices (IUD) and contraceptive implants) are safe, highly effective, and satisfactory options available to women who have been appropriately counseled [3]. Yet, multiple barriers to utilization have been identified [4–6] including hesitation from providers on ‘early’ LARC removal [7–9], delay in placement [10], and variations in available individual health coverage [11–12]. As of September 2010, the Affordable Care Act (ACA) requires many insurance plans to provide in-network coverage without cost sharing of certain clinical preventive services including all FDA-approved contraceptive methods [13]; however, additional requirements for reimbursement of services in individual state-level policies, exist. For instance, health plans or issuers of plans may use reasonable medical management techniques to control cost by imposing cost sharing when equivalent branded drugs are used [13]. Barriers to LARC removal and reinsertion access may also occur due to imposing prior authorization and step therapy, approval for medically necessary procedures only [9], or other non-medical reasons [11]. Therefore, it is important to understand how reimbursement policies for LARC devices, specifically for removal and reinsertion, affect health services delivery at the population level. This review summarizes language in state-level reimbursement policies on LARC removal and reinsertion, and language on reimbursement requirements.

## Materials and methods

*S*tudy authors conducted a systematic, web-based review of publicly available state-level documents from October 2017 to May 2018. Detailed search terms, data abstraction process, and methodology are described elsewhere [14–15]. Briefly, reimbursement policies (e.g., Medicaid Bulletin, Family Planning Waiver, State Plan Amendment) authored by the state or an entity with authority to create billing policies, were categorized as ‘State issued’. We used the term ‘Health Plan’, to categorize polices (e.g., Provider Manual and Insurance Manual) authored from a health plan with authority from the state to bill for services. When developing the definition of state-based reimbursement policies for LARC removal or reinsertion, study authors reviewed language in all documents that referred to or detailed reimbursement for LARC. If the word ‘removal’ or ‘reinsertion’ was included in the policy language or if the policy contained International Classification of Diseases codes (e.g., Z30.46, Z30.433) or Current Procedural Terminology (e.g., 11982, 11983, 58301) representing removal or reinsertion of a LARC device or Healthcare Common Procedure Coding System codes (e.g., J7296, J7297, J7300, J7307), [16] the state was categorized as having reimbursement policies for LARC removal or reinsertion.

Likewise, a state was categorized as having reimbursement language for reinsertion if the language included words such as ‘replacement’ ‘maintain’ and/or ‘re-implanted’ when describing LARC services or reimbursement policies.

We categorized reimbursement requirements for removal or reinsertion into ‘not specified’ if policies did not specify reimbursement requirements for LARC removal or reinsertion, ‘no requirement for provision of services’ if the language prohibited limitations on removal or reinsertion services, and ‘specified’ if specific requirements were mentioned. Among policies with specific language, we categorized requirements into the following groupings: *Coverage-related requirements*―represented policy language that limits reimbursement to preferred in-network providers or by other stipulations in the members’ benefit. *Step-therapy related requirements*―allowed for reimbursement only after a therapeutic equivalency device has been used. *Time-related requirements*―limited reimbursement to mandated periods of effectiveness (e.g., 3 years), or required minimum time allotment prior to a device’s removal or reinsertion (e.g., 6 months). *Diagnosis-related requirements*―limited reimbursement to when the removal or reinsertion was needed secondary to the presence of a medical condition (e.g., bleeding issues, infection) or when the patient was treated for an unrelated diagnosis or for a visit not coded as a family planning visit. Lastly, *same-day related requirements*―represented language that limits reimbursement to same day visits.

We used descriptive statistics to analyze the abstracted information. This study was determined to be public health practice and, therefore, did not require Institutional Review Board approval at the Centers for Disease Control and Prevention or the University of Illinois at Chicago.

## Results

Twenty-six (52%) of the 50 states had publicly available policies that addressed reimbursement for LARC removal or reinsertion Table 1.

**Table 1:** Publicly Available Reimbursement Policies on LARC Removal Or Reinsertion By Policy Type and Source for All States, 2017–2018. (N=50)

| States | Policy Characteristics |  |  |  |
| --- | --- | --- | --- | --- |
|  | Publicly Available Policy<sup>a</sup> | Policy Types |  | Policy Source |
|  |  | State Issued<sup>b</sup> | Health plan<sup>c</sup> |  |
| Alabama | Yes | Yes | Yes | Medicaid Guidance & Health Plan Benefit Guide |
| Alaska | — | — | — | — |
| Arizona | — | — | — | — |
| Arkansas | Yes | Not available<sup>d</sup> | Yes | Health Plan Benefit Guide |
| California | Yes | Not available | Yes | Health Plan Benefit Guide |
| Colorado | — | — | — | — |
| Connecticut | — | — | — | — |
| Delaware | — | — | — | — |
| Florida | — | — | — | — |
| Georgia | — | — | — | — |
| Hawaii | Yes | Not available | Yes | Health Plan Benefit Guide |
| Idaho | Yes | Yes | Not available | Medicaid Guidance |
| Illinois | Yes | Yes | Yes | Statutory Provision & Health Plan Benefit Guide |
| Indiana | — | — | — | — |
| Iowa | — | — | — | — |
| Kansas | Yes | Yes | Yes | State Plan Amendment & Health Plan Benefit Guide |
| Kentucky | Yes | Not available | Yes | Health Plan Benefit Guide |
| Louisiana | — | — | — | — |
| Maine | Yes | Yes | Not available | Medicaid Guidance |
| Maryland | — | — | — | — |
| Massachusetts | Yes | Yes | Not available | Medicaid Guidance |
| Michigan | — | — | — | — |
| Minnesota | Yes | Not available | Yes | Health Plan Benefit Guide |
| Mississippi | — | — | — | — |
| Missouri | Yes | Yes | Not available | Medicaid Guidance |
| Montana | Yes | Yes | Not available | Medicaid Guidance |
| Nebraska | Yes | Yes | Not available | Medicaid Guidance & Title X |
| Nevada | Yes | Yes | Not available | Medicaid Guidance |
| New Hampshire | Yes | Yes | Not available | Medicaid Guidance |
| New Jersey | Yes | Not available | Yes | Health Plan Benefit Guide |
| New Mexico | Yes | Yes | Not available | Medicaid Guidance |
| New York | — | — | — | — |
| North Carolina | Yes | Yes | Yes | Statutory Provision & Health Plan Benefit Guide |
| North Dakota | Yes | Yes | Not available | Medicaid Guidance |
| Ohio | — | — | — | — |
| Oklahoma | Yes | Yes | Not available | State Plan Amendment |
| Oregon | — | — | — | — |
| Pennsylvania | — | — | — | — |
| Rhode Island | — | — | — | — |
| South Carolina | — | — | — | — |
| South Dakota | Yes | Yes | Not available | Medicaid Guidance |
| Tennessee | — | — | — | — |
| Texas | — | — | — | — |
| Utah | Yes | Not available | Yes | Health Plan Benefit Guide |
| Vermont | — | — | — | — |
| Virginia | — | — | — | — |
| Washington | Yes | Yes | Yes | Medicaid Guidance & Health Plan Benefit Guide |
| West Virginia | Yes | Not available | Yes | Health Plan Benefit Guide |
| Wisconsin | Yes | Yes | Yes | Statutory Provision & Health Plan Benefit Guide |
| Wyoming | — | — | — | — |

LARC, Long-acting reversible contraception
<sup>a</sup> The dashes in these columns represent states that **did not** have a publicly available policy.
<sup>b</sup> State issued policy type represents reimbursement policies authored by the state or an entity with authority to create billing policies such as Medicaid, a Statute, or a State Plan Amendment.
<sup>c</sup> Health plan policy type represents reimbursement policies authored by a health plan, with authority from the state, to bill for services within the state.
<sup>d</sup> ‘Not available’ represents policies that did not specify a statewide or a health plan policy type.

While all 26 states included language in policies that addressed reimbursement for LARC removal, only 14 policies included language to address reinsertion Table 2.

**Table 2:** Summary of Reimbursement Policies and Requirements on LARC Removal or Reinsertion Among States with Publicly Available Policies, 2017–2018 (N=26)

| **State with Publicly Available Policies** | **Reimbursement Policy for LARC Removal** | **Requirement for Reimbursing LARC Removal <sup>a</sup>** | **Reimbursement Policy for LARC Reinsertion** | **Requirement for Reimbursing LARC Reinsertion** |
| --- | --- | --- | --- | --- |
| Alabama | Yes | Not specified | — <sup>b</sup> | — |
| Arkansas | Yes | Not specified | — | — |
| California | Yes | Coverage-related requirements<br>Step-therapy related requirements | — | — |
| Hawaii | Yes | Time-related requirements | — | — |
| Idaho | Yes | Diagnosis-related requirements | — | — |
| Illinois | Yes | No restriction for provision of services | Yes | No restriction for provision of services |
| Kansas | Yes | Not specified | — | — |
| Kentucky | Yes | Not specified | Yes | Time-related requirements |
| Maine | Yes | Not specified | — | — |
| Massachusetts | Yes | Not specified | Yes | Not specified |
| Minnesota | Yes | Not specified | Yes | Not specified |
| Missouri | Yes | Not specified | Yes | Not specified |
| Montana | Yes | Not specified | Yes | Not specified |
| Nebraska | Yes | Same-day related requirements | Yes | Same-day related requirements |
| Nevada | Yes | Step-therapy related requirements | Yes | Step-therapy related requirements |
| New Hampshire | Yes | Not specified | Yes | Not specified |
| New Jersey | Yes | Coverage-related requirements | Yes | Coverage-related requirements |
| New Mexico | Yes | Not specified | — | — |
| North Carolina | Yes | No restriction for provision of services | — | — |
| North Dakota | Yes | Diagnosis-related requirements | — | — |
| Oklahoma | Yes | No restriction for provision of services | Yes | Time-related requirements |
| South Dakota | Yes | Time-related requirements | — | — |
| Utah | Yes | Not specified | — | — |
| Washington | Yes | Not specified | Yes | Not specified |
| West Virginia | Yes | Not specified | Yes | Time-related requirements |
| Wisconsin | Yes | Not specified | Yes | Not specified |

LARC, Long-acting reversible contraception
<sup>a</sup> *Not specified* represents policies that did not specify reimbursement requirements for LARC removal or reinsertion. *Coverage-related requirements* represent policy language that limits the reimbursement of LARC removal or reinsertion to preferred in-network providers or by other stipulations in the members' benefit coverage. *Step-therapy related requirements* represent policy language that allows for removal or reinsertion of LARC reimbursement only after a therapeutic equivalency device has been used. *Time-related requirements* represent policy language that limits the reimbursement of LARC removal or reinsertion to mandated periods of effectiveness (e.g., 3 years), or requires minimum time allotment prior to a device's removal or reinsertion (e.g., 6 months). *Diagnosis-related requirements* represent policy language that limits the reimbursement of LARC removal or reinsertion to when the removal or reinsertion was needed secondary to the presence of a medical condition (e.g., bleeding issues, infection) or when the patient was treated for an unrelated diagnosis or for a visit not coded as a family planning visit. *No restriction for provision of services* represents policies where the provided language specifically prohibits restrictions on removal or reinsertion services. *Same-day related requirements* represent policy language that limits reimbursement and no cost sharing, for the removal or reinsertion of LARC, to same day visits.
<sup>b</sup> The dashes in these columns represent policies that **did not** include language on reinsertion or its reimbursement.

Three policies included language that specified no restrictions be placed on reimbursement for removal and one state specified no restrictions on reimbursement for reinsertion. The most common type of reimbursement requirement was time-related (n=5); the least common was same-day related requirements (n=1). Table 2.

## Discussion and conclusions

We found that more than a quarter of states had policy language on reimbursement for LARC removal, while fewer addressed reimbursement for reinsertion. Only three states had policy language indicating no reimbursement requirement for provision of services. Most states with a publicly available reimbursement policy for LARC removal or reinsertion were Medicaid policies, with few states’ Health Plans polices publicly available for review. The public availability of more Medicaid policies likely reflects the efforts undertaken by the Centers for Medicare and Medicaid Services (CMS)/Center for Medicaid and Children’s Health Insurance Program Services (CMCS) who, in 2014, launched the Maternal and Infant Health Initiative with the primary goal of increasing access and use of effective contraceptives including LARC [17] though use among Medicaid recipients varies by state [17–18].

One potential reason for requirements for reimbursement could be concerns that ‘early’ removal would be costly [19–20]. However, LARC devices are cost neutral as early as three months post insertion, prior to full duration of effectiveness, when compared with short-acting reversible contraception options (i.e., patches, rings, oral contraceptive pills and injections) or no method use at all [21]. This finding of cost neutrality is still present even when the cost implications of removing the device before the end of its effective date is included [21].

Our findings of state-level variation in LARC removal and reinsertion reimbursement policies is consistent with existing literature demonstrating variation in LARC access policies. [9,11,14,15, 22]. Specific reimbursement practices may present barriers for LARC removal or reinsertion. For women in states with policies that include reimbursement requirements, such as diagnosis and time-related requirements, preferences for LARC maybe impacted if women lack assurance that removal will be covered [9,23]. Moreover, access to LARC removal or reinsertion without restrictions is vitally important, particularly for populations who have experienced restraint of reproductive autonomy (e.g., American Indian/Alaskan Native people, Black people, people with disabilities, people experiencing poverty and people who are incarcerated or detained) [24–30].

Recognizing these concerns, national clinical organizations encourage patient-centered counseling based on individual patient contraceptive preferences, needs, and values, thus ensuring that patient values guide all clinical decisions [31]. Similarly, the American College of Obstetricians and Gynecologists recommends a reproductive justice framework be employed during contraceptive counseling which entails shared decision-making with the patient and provision of information on the benefits and risks of all contraceptive methods with the avoidance of potential coercion [32]. Recently, a multidisciplinary group of experts developed a Reproductive and Sexual Health Equity framework; a key principle is the concept of honoring bodily autonomy, emphasizing ongoing difficulties women have accessing LARC removal [33].

Several limitations exist in interpreting our findings. First, we did not contact all states to verify their reimbursement policies on removal or reinsertion of LARC. Second, we only included publicly available policies, potentially missing any new, non-publicly available or unpublished policies. Third, while our reviewed focused on reimbursement policies and its effect on LARC access, numerous other barriers such as lack of provider knowledge [34], blocked time for provider training [35], credentialing gaps [36], and myths and misinformation from patients [32] may contribute to the ability of women to access LARC removal and/or reinsertion. Fourth, since the data collection timeframe, some state policies may have been reviewed or amended, potentially affecting our categorization of policy language. However, amendment of state policies may require multiple annual policy cycles depending on whether the policy is a law, regulation, standard, or protocol [17].

Given that reimbursement policies can influence service delivery [37], review of language may identify administrative, financial, or medical barriers to reimbursement for LARC removal or reinsertion [9,30,38]. For example, states could consider including reimbursement language that allows providers to bill per-service rather than per-visit, allowing insertion, removal, or reinsertion of a LARC during a single clinical encounter [39], if desired by the woman. Lastly, standardizing training of family medicine [35] or primary care residents [36] on insertion and removal with expanded reimbursement could increases timely access to LARCs.

LARC removal and reinsertion are important aspects of contraception access. Reimbursement requirements may restrict contraceptive access. Removal of barriers to these services supports both the ability of providers to offer comprehensive contraceptive services, and patient reproductive autonomy.

## Data Availability

Data are all publicly available. Detailed search terms, data abstraction process, and methodology are described and referenced in the methodology section.

## Acknowledgements

None.

## Notes

### Competing Interest Statement

The authors have declared no competing interest.

### Funding Statement

The author(s) received no specific funding for this work.

### Author Declarations

This study was determined to be public health practice and, therefore, did not require Institutional Review Board approval at the Centers for Disease Control and Prevention or the University of Illinois at Chicago.

